# Up-regulated tumor intrinsic growth potential and decreased immune function orchestrate the evolution of lung adenocarcinoma

**DOI:** 10.1101/2022.01.01.22268606

**Authors:** Yue Zhao, Jun Shang, Jian Gao, Han Han, Zhendong Gao, Yueren Yan, Qiang Zheng, Ting Ye, Fangqiu Fu, Chaoqiang Deng, Zelin Ma, Yang Zhang, Difan Zheng, Shanbo Zheng, Yuan Li, Zhiwei Cao, Leming Shi, Haiquan Chen

## Abstract

**Introduction:** Lung adenocarcinoma is the most common pathological subtype of lung cancer. Precursors of lung adenocarcinoma, namely adenocarcinoma *in situ* and minimally invasive adenocarcinoma, have a superb 5-year survival rate after surgical resection. A deeper understanding of the key genetic changes driving the progression of lung adenocarcinoma is needed.

**Methods:** In this study, we performed whole-exome sequencing and RNA-sequencing on surgically resected 24 AIS, 74 MIA, 99 LUAD specimens and their adjacent paired normal tissues. Radiological, clinical, and pathological characteristics were recorded. Gene expression patterns were identified to find key pathways driving the progression of lung adenocarcinoma. Furthermore, genomic alterations and differential expression analyses were performed to compare tumors with different radiological manifestations. Finally, a progressive index was developed to quantitatively measure the level of imbalance between tumor intrinsic growth potential and immune microenvironment.

**Results:** 12 patterns of gene expression were identified. Pathways associated with tumor growth and metastasis were found to be up-regulated as tumors progressed, while pathways associated with immune function were found to be down-regulated. Deconvolution of RNA-seq data also showed a decrease of CD8+ T cells and an increase of Tregs as the tumors progressed. Furthermore, tumors with more solid components on CT scan had a higher mutation frequency of tumor suppressor genes, higher tumor mutation burden and higher frequency of somatic copy number alterations. Finally, tumor progressive index demonstrated an increasing trend with the progression of lung adenocarcinoma.

**Discussion:** Our results reveal the imbalance of tumor intrinsic factors and immune function orchestrate the progression of lung adenocarcinoma.

## Introduction

Lung cancer is one of the deadliest disease worldwide, with a 5-year survival of only ∼19%^1^. Lung adenocarcinoma (LUAD) is the most common pathological subtype. Pre-invasive stages of lung adenocarcinoma, namely adenocarcinoma *in situ* (AIS) and minimally invasive adenocarcinoma (MIA), have a nearly 100% 5-year survival rate after complete surgery^2,3^. Although there have been studies on genomic and immune profiling of patients with AIS, MIA and LUAD, there lack a systematic study focusing on key molecular events that drive the evolution of lung adenocarcinoma^4–8^.

With the development of thoracic computed tomography (CT) scanning and the application of low-dose CT screening, an increasing number of small pulmonary nodules, especially subsolid nodules have been detected^9–12^. The prognostic impact of solid components for lung adenocarcinomas presented as ground-glass opacities (GGO) on CT scanning has been under extensive investigation, where tumors manifesting as GGOs were found to have indolent clinical course^13,14^ Our two previous studies further provided evidence on the prognostic value of lung adenocarcinomas manifesting as GGOs on CT scan^15,16^. However, there still lack comprehensive genomic and transcriptomic studies comparing the differences between lung adenocarcinomas having GGO components and their counterparts not having GGO components on CT scan.

In this study, we performed whole-exome sequencing and RNA-sequencing on 197 surgically resected lung adenocarcinomas and divided them by pathological characteristics and radiological manifestations, aiming to find key genetic factors that drive the evolution of lung adenocarcinoma. 12 expression patterns were identified based on expression profiles, and pathway analysis was performed to reveal the biological functions for each pattern. Tumor intrinsic growth potential and immune microenvironment were assessed, and immune cell infiltration was calculated using transcriptomic data. Finally, a tumor progressive index was developed to provide a quantitative measurement of the level of unbalance between tumor intrinsic growth potential and immune function.

## Methods

### Study cohort

197 lung adenocarcinoma patients who underwent surgery between September 2011 and May 2016 at the Department of Thoracic Surgery, Fudan University Shanghai Cancer Center were retrospectively included in this study (figure 1A). None of the patients received neoadjuvant therapy. This study was approved by the Committee for Ethical Review of Research (Fudan University Shanghai Cancer Center Institutional Review Board No. 090977-1). Informed consents of all patients for donating their samples to the tissue bank of Fudan University Shanghai Cancer Center were obtained from patients themselves or their relatives. Radiological, histological evaluation and Follow-up protocol are described in detail in supplementary methods.

**Figure 1.**
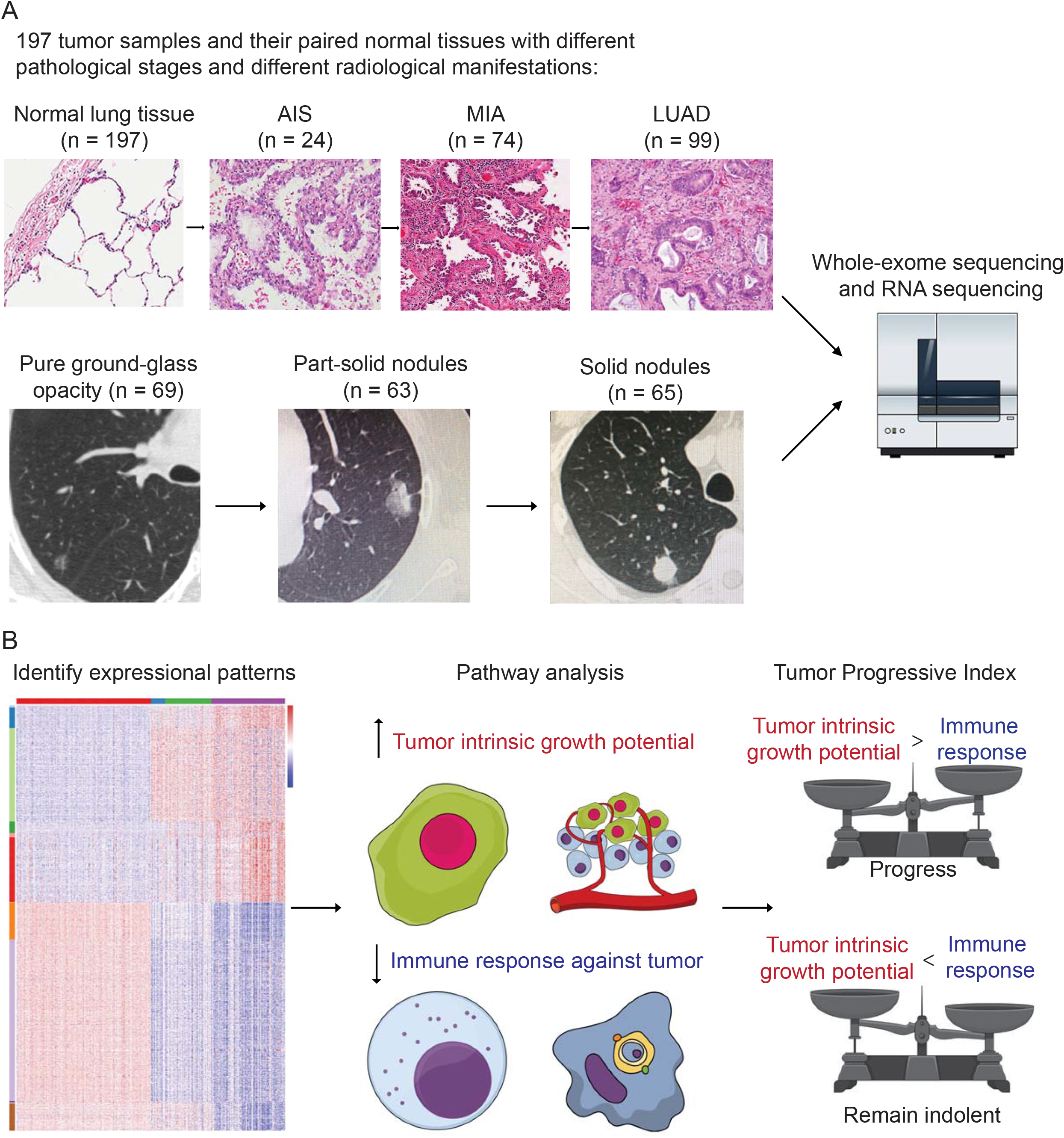
Study design. A) a total number of 197 tumor samples, including 24 adenocarcinoma *in situ* (AIS), 74 minimally invasive adenocarcinoma (MIA), 99 lung adenocarcinoma (LUAD) and their paired adjacent normal lung tissue underwent whole-exome sequencing and RNA-sequencing. Genomic and transcriptomic data were analyzed and compared among different groups. Samples were further divided into 3 groups according to their radiological manifestations: 69 pure ground-glass opacities (GGOs), 63 subsolid nodules and 65 solid nodules. B) Identification of 12 expressional patterns and development of tumor progressive index based on genomic and transcriptomic data.

### Radiological and histological evaluation

Whole lung CT scans were performed on each patient included before surgery as previously described^15^. For each nodule, the maximum diameter of both the entire nodule and solid component on the single largest axial dimension was recorded on lung window. Pulmonary nodules were further divided into 3 groups: pure GGOs, where there was no solid component in one pulmonary nodule; subsolid nodules, where both solid and GGO components existed in one pulmonary nodule; solid nodules, where the pulmonary nodule contained only solid component (figure 1A). CT scans were reviewed by two radiologists independently, and interobserver and intraobserver agreements were measured to quantify the reproducibility and accuracy between the two radiologists as previously described^15^.

Intraoperative frozen section diagnosis was made after the tumor was resected, and postoperative diagnosis was made after surgery by two independent pathologists. According to the IASLC/ATS/ERS classification, tumors were classified as adenocarcinoma *in situ* (AIS), minimally invasive adenocarcinoma (MIA) or invasive lung adenocarcinoma (LUAD) based on their histological presentations. Invasive lung adenocarcinoma subtypes were further analyzed in a semi-quantitative manner, where the components of different subtypes (lepidic, acinar, papillary, micropapillary, solid and invasive mucinous) were recorded in 5% increments. The predominant subtype was the one with the largest percentage (not necessarily 50% or higher)^17^. Pathological stage of disease was determined according to the 8th TNM staging system.

### Follow-up protocol

Patients were followed up regularly after surgery as we previously described^15^. Briefly, patients were followed up every 3 months after surgery for the first 2 years, where physical examination, chest CT scans and abdominal ultrasonography were performed every 3 to 6 months. The follow-up interval was changed to every 6 months for the third year and once a year from then on. Brain CT or magnetic resonance imaging (MRI) and bone scintigraphy were performed every 6 months for patients with invasive adenocarcinoma in the first 3 years. In addition, positron emission tomography-CT (PET-CT) scans was performed if necessary. Recurrence-free survival (RFS) was defined as the time between surgery and first recurrence or last follow-up. Patients with no recurrence but died from other causes were censored on that date. Overall survival (OS) was defined as the time between surgery and death or last follow-up.

### Whole-exome sequencing

Genomic DNA from tumors and paired adjacent normal tissues were extracted and prepared using the QIAamp DNA Mini Kit (Qiagen, Germany) following the manufacturer’s instructions. Exon libraries were constructed using the SureSelect XT Target Enrichment System. A total amount of 1-3 μg genomic DNA for each samples was fragmented into an average size of ∼200 bp. DNA was hybridized, captured and amplified using SureSelect XT reagents and protocols to generate indexed, target-enriched library amplicons. Constructed libraries were then sequenced on the Illumina HiSeq X Ten platform and 150 bp paired-end reads were generated.

### Alignment, mutation calling and somatic copy number alteration calling

Sequence reads from the exome capture libraries were aligned to the reference human genome (hg19) using BWA-MEM^18^. Picard tools was then used for marking PCR duplicated and the Genome Analysis Toolkit (GATK) was used to perform base quality recalibration and local indel re-alignments^19^. Single nucleotide variants (SNVs) were called using MuTect and MuTect2^20^. Indels were called using MuTect2 and Strelka v2.0.13^21^. Variantes were filtered if called by only one tool. After the variants were called, Oncotator v1.9.1 was used for annotating somatic mutations^22^, and significantly mutated genes were identified using MutSig2CV^23^. Tumor mutation burden (TMB) was calculated as the total number of nonsynonymous SNVs and indels per sample divided by 30, given the total coverage of ∼30 Mb. CNVkit v0.9.7 with default parameters was used to perform somatic copy number alteration (SCNA) analysis for alignment reads^24^. Amplification and deletion peaks were identified using GISTIC2.0 from segment files^25^. Amplification and deletion thresholds were set 0.1 and −0.1, respectively. Frequency distribution of amplification and deletion was shown using R package copynumber v1.26.0^26^.

### RNA-sequencing and calculation of expression

Total RNA from tumors and paired adjacent normal tissues was extracted and prepared using NucleoZOL (Macherey-Nagel, Germany) and NucleoSpin RNA Set for NucleoZOL (Macherey-Nagel, Germany) following the manufacturer’s instructions. A total amount of 3 μg RNA per sample was used as initial material for RNA sample preparations. Ribosomal RNA was removed using Epicenter Ribo-Zero Gold Kits (Epicenter, USA). Sequencing libraries were generated using NEBNext Ultra Directional RNA Library Prep Kit for Illumina (NEB, Ipswich, USA) according to the manufacturer’s instructions. Libraries were then sequenced on the Illumina HiSeq X Ten platform and 150 bp paired-end reads were generated. As a quality-control step, only those samples with RNA integrity number (RIN) >= 5.0 were included in this study.

After the data were obtained, RNA-seq reads were aligned to the reference human genome (hg19) using STAR v2.5.3^27^. Expression values were normalized to the transcripts per million (TPM) estimates using RSEM v1.3.0^28^.

### Identification of gene expression patterns

150 samples with RIN >= 5.0 were divided into 4 groups: normal (n = 150), AIS (n = 16), MIA (n = 52) and LUAD (n = 82). Analysis of variance (ANOVA) was used to determine differentially expressed genes. First, paired comparison was performed between two adjacent groups (normal vs AIS, AIS vs MIA and MIA vs LUAD), then genes with p < 0.0001 and |fold change| >= 2 in at least 1 of 3 comparisons were considered significant and put into downstream analyses. 12 patterns of gene expression were then identified based on their trends of expression between each two adjacent groups in the study cohort (up trend, no significant change or down trend, figure 1B and figure 2). Finally, KOBAS was used to perform KEGG pathway enrichment analysis for the patterns identified in the study cohort^29^.

**Figure 2.**
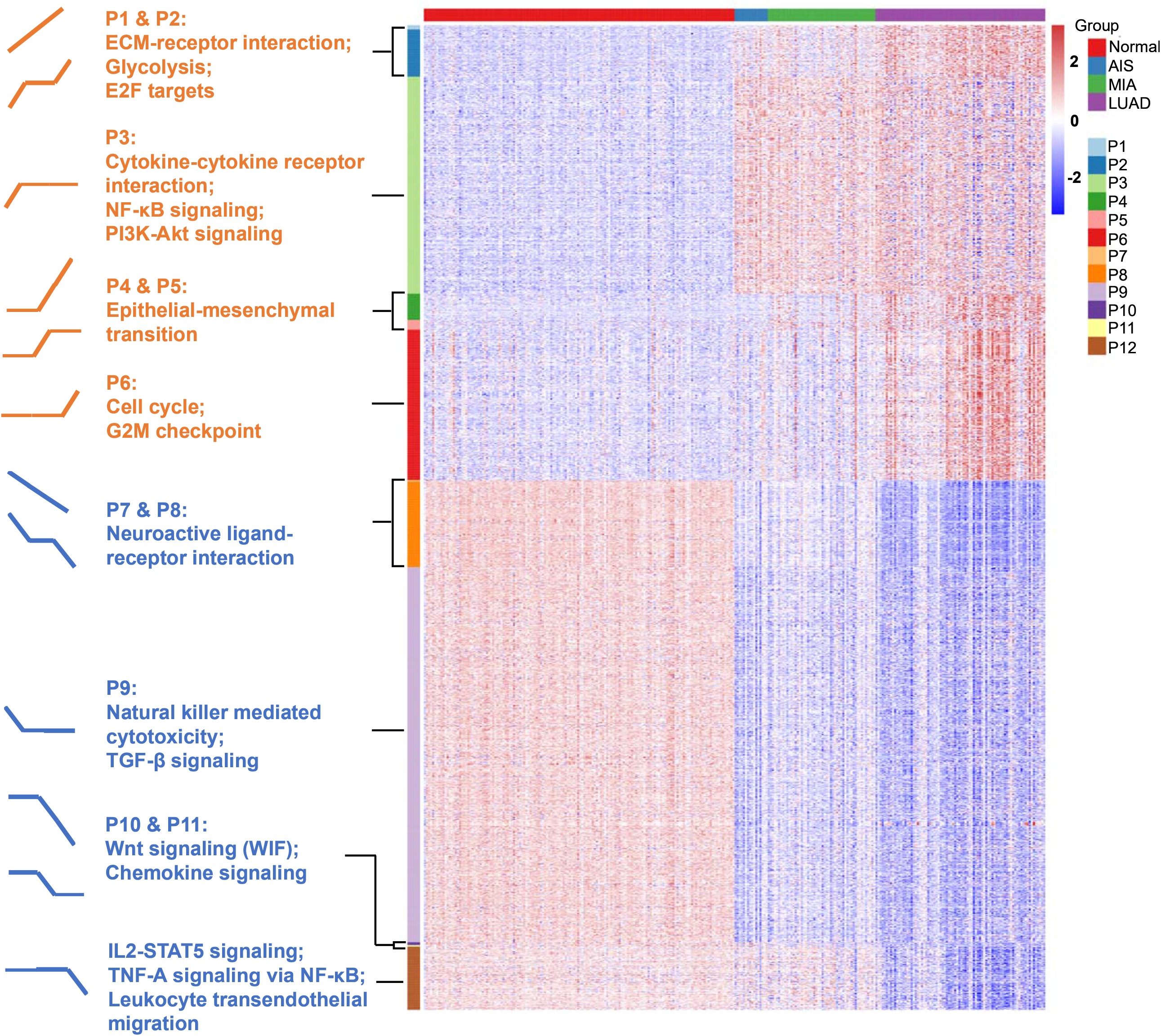
Identification of 12 expression patterns. Pattern 1 (9 genes): increase from normal to AIS, from AIS to MIA and from MIA to LUAD; pattern 2 (97 genes): increase from normal to AIS and from MIA to LUAD, no change from AIS to MIA; pattern 3 (446 genes): increase from normal to AIS alone; pattern 4 (54 genes): no change from normal to AIS, increase from AIS to MIA and from MIA to LUAD; pattern 5 (20 genes): increase from AIS to MIA alone; pattern 6 (309 genes): increase from MIA to LUAD alone; pattern 7 (3 genes): decrease from normal to AIS, from AIS to MIA and from MIA to LUAD; pattern 8 (175 genes): decrease from normal to AIS and from MIA to LUAD, no change from AIS to MIA; pattern 9 (771 genes): decrease from normal to AIS alone; pattern 10 (6 genes): no change from normal to AIS, decrease from AIS to MIA and from MIA to LUAD; pattern 11 (3 genes): decrease from AIS to MIA alone; pattern 12 (130 genes): decrease from MIA to LUAD alone. Gene set enrichment analysis (GSEA) was performed to assess the functional significance for those patterns.

### Differential expression analysis and gene set enrichment analysis

Differential expression analysis was performed using DESeq2^30^. False discovery rate (FDR) was controlled using the Benjamini-Hochberg method. Genes with log2-fold change ≥ 1 or ≤ −1, and adjusted p value < 0.05 were considered differentially expressed between two groups. Significantly differentially expressed genes were then included in the downstream gene set enrichment analysis (GSEA). GSEA was performed using the GSEA software (version 3.0)^31,32^. Briefly, normalized enrichment scores (NES) and p values were calculated to sort the pathways enriched in each group. Cancer hallmark pathways were used for this analysis, and gene set permutations were performed 1000 times for each analysis. Enrichment maps were generated to visualize the results.

### Development of tumor progressive index

Tumor progressive index was developed for quantitatively measuring the level of imbalance between tumor intrinsic growth potential and immune response. To minimize the possible confounding effect introduced by genes with low expression, we first filtered out genes with mean TPM < 1.0 across all samples. Next, genes for calculating the tumor index were selected from Pattern 1 where there was a significant increase between each two adjacent groups. Genes for calculating the immune index were selected from Pattern 8 where there was a significant decrease from normal tissue to AIS and from MIA to LUAD. Those genes that were both selected form Pattern 8 and were in an *a priori* immune-related gene list containing 730 genes was used to calculate the immune index^7^. For both tumor index and immune index, expression for those genes across all samples were first transformed to Z-scores, then for each sample, tumor index was calculated as the mean value of Z-scores for the tumor index genes, while immune index was calculated as the mean value of Z-scores for immune index genes. Finally, tumor progressive index was calculated as the subtraction of tumor index by immune index:

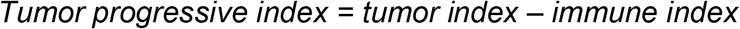

### Statistical analysis

Clinical and pathological characteristics were recorded and compared among three groups. Pearson χ^2^ test and Fisher’s exact test were used to compare categorical variables wherever applicable. Non-parametric Wilcoxon signed-rank test was used to compare medians of groups of continuous variables. Kaplan-Meier survival curves and log-rank p values were calculated for patients’ RFS and OS. All statistical analyses and graphing work were performed using R (version 3.6.0, R Foundation for Statistical Computing, Vienna, Austria). Two-tailed p < 0.05 was considered significant for all statistical analyses unless stated otherwise.

## Results

### Association between pathological stage and radiological manifestations

Tumor samples were divided into 3 groups based on their pathological findings: 24 AIS, 74 MIA and 99 LUAD, and divided into 3 groups based on their radiological manifestations: 69 pure GGOs, 63 subsolid nodules and 65 solid nodules (figure 1A). Clinical and pathological characteristics, including sex, smoking status, tumor location, pathology and adenocarcinoma subtypes were compared (table 1). Of note, we found that there was an association between radiological and pathological presentations. 66 of 69 (95.7%) of pulmonary nodules manifesting as pure GGOs on CT scan were either adenocarcinoma *in situ* (AIS) or minimally invasive adenocarcinoma (MIA), while for pulmonary nodules manifesting as pure solid nodules, only 1 out of 65 (1.5%) was either AIS or MIA (MIA in this case). For pulmonary nodules manifesting as subsolid nodules on CT scan, 31 out of 63 (49.2%) were either AIS or MIA, and 32 (50.8%) were invasive adenocarcinoma. Compared to male patients, female patients had significantly higher frequency of pure GGO lesions (p = 0.025). For predominant adenocarcinoma subtypes, lepidic subtype was significantly enriched in the pure GGO group (p < 0.001), while papillary subtype was significantly enriched in the pure solid group (p = 0.004). Solid subtype was only found predominant in the pure solid group. Moreover, for presenting subtypes, lepidic subtype was significantly enriched in the pure GGO group (p = 0.001), and solid subtype was significantly enriched in the pure solid group (p = 0.014). Micropapillary subtype was also more likely to be found in the pure solid group, though without a significant difference (pure GGO vs subsolid vs solid: 0.0% vs 3.1% vs 9.4%, p = 0.472). Taken together, these results demonstrated there was a link between radiological and pathological findings, therefore, radiological manifestations can, at least partly, help predict the invasiveness of lung adenocarcinoma.

**Table 1.** Clinical and pathological characteristics of the study cohort (n = 197). Abbreviations: AIS, adenocarcinoma *in situ*; GGO, ground-glass opacity; IAD, invasive adenocarcinoma; IMA, invasive mucinous adenocarcinoma; MIA, minimally invasive adenocarcinoma; LLL, left lower lobe; LUL, left upper lobe; RLL, right lower lobe; RML, right middle lobe; RUL, right upper lobe.

|  | Pure GGO<br>(n = 69) | Subsolid<br>(n = 63) | Solid<br>(n = 65) | P-value |
| --- | --- | --- | --- | --- |
| Sex |  |  |  | 0.025 |
| Female | 48 (69.6%) | 41 (65.1%) | 31 (47.7%) |  |
| Male | 21 (30.4%) | 22 (34.9%) | 34 (52.3%) |  |
| Smoking status |  |  |  | 0.212 |
| Former/current | 16 (23.2%) | 15 (23.8%) | 23 (35.4%) |  |
| Never | 53 (76.8%) | 48 (76.2%) | 42 (64.6%) |  |
| Tumor location |  |  |  | 0.661 |
| LUL | 18 (26.1%) | 13 (20.6%) | 11 (16.9%) |  |
| LLL | 7 (10.1%) | 6 (9.5%) | 10 (15.4%) |  |
| RUL | 26 (37.7%) | 31 (49.2%) | 23 (35.4%) |  |
| RML | 7 (10.1%) | 5 (7.9%) | 8 (12.3%) |  |
| RLL | 11 (15.9%) | 8 (12.7%) | 13 (20.0%) |  |
| Pathology |  |  |  | < 0.001 |
| AIS/MIA | 66 (95.7%) | 31 (49.2%) | 1 (1.5%) |  |
| IAD | 3 (4.3%) | 32 (50.8%) | 64 (98.5%) |  |
| Predominant subtype |  |  |  |  |
| Lepidic | 2 (66.7%) | 9 (28.1%) | 3 (4.7%) | < 0.001 |
| Acinar | 1 (33.3%) | 17 (53.1%) | 40 (62.5%) | 0.453 |
| Papillary | 0 (0.0%) | 6 (9.4%) | 10 (15.6%) | 0.004 |
| Micropapillary | 0 (0.0%) | 0 (0.0%) | 0 (0.0%) | - |
| Solid | 0 (0.0%) | 0 (0.0%) | 9 (14.1%) | 0.067 |
| IMA | 0 (0.0%) | 0 (0.0%) | 2 (3.1%) | 0.123 |
| Presenting subtype |  |  |  |  |
| Lepidic | 2 (66.7%) | 12 (37.5%) | 6 (9.4%) | 0.001 |
| Acinar | 2 (66.7%) | 24 (75.0%) | 50 (78.1%) | 0.863 |
| Papillary | 0 (0.0%) | 9 (28.1%) | 22 (34.4%) | 0.407 |
| Micropapillary | 0 (0.0%) | 1 (3.1%) | 6 (9.4%) | 0.472 |
| Solid | 0 (0.0%) | 1 (3.1%) | 17 (26.6%) | 0.014 |
| IMA | 0 (0.0%) | 0 (0.0%) | 2 (3.1%) | 0.572 |

### 12 expression patterns were identified as tumors progress

One aim of this study is to find key genetic factors that drive the evolution of lung adenocarcinoma. 150 samples with RNA integrity number (RIN) >= 5.0 were included in this part. We developed an ANOVA model with pathological stage (normal tissue, AIS, MIA and LUAD) as a factor and patient as a random effect. 2023 genes with p < 0.0001 and |fold change| > 2 in pairwise comparisons of two adjacent groups of patients (normal tissue vs AIS, AIS vs MIA, and MIA vs LUAD, respectively) were divided into 12 following patterns: Pattern 1 (9 genes): increase from normal to AIS, from AIS to MIA and from MIA to LUAD; pattern 2 (97 genes): increase from normal to AIS and from MIA to LUAD, no change from AIS to MIA; pattern 3 (446 genes): increase from normal to AIS alone; pattern 4 (54 genes): no change from normal to AIS, increase from AIS to MIA and from MIA to LUAD; pattern 5 (20 genes): increase from AIS to MIA alone; pattern 6 (309 genes): increase from MIA to LUAD alone; pattern 7 (3 genes): decrease from normal to AIS, from AIS to MIA and from MIA to LUAD; pattern 8 (175 genes): decrease from normal to AIS and from MIA to LUAD, no change from AIS to MIA; pattern 9 (771 genes): decrease from normal to AIS alone; pattern 10 (6 genes): no change from normal to AIS, decrease from AIS to MIA and from MIA to LUAD; pattern 11 (3 genes): decrease from AIS to MIA alone; pattern 12 (130 genes): decrease from MIA to LUAD alone (figure 2 and supplementary table 1). Pathway enrichment analysis showed that pathways associated with tumor invasiveness and cell growth were enriched in up-trend patterns (patterns 1 to 6). Of note, PI3K-AKT signaling pathway and NF-κB signaling pathway were up-regulated in AIS compared with normal tissue, suggesting malignant behaviors could exist in as early as AIS. Epithelial-mesenchymal transition, which was highly associated with tumor metastasis, was also found to be increased from AIS to MIA, suggesting an acquisition of metastatic potential as invasive subtypes of lung adenocarcinoma emerge^33^. Moreover, cell cycle was found to be increased from MIA to LUAD, consistent with the fact that AIS/MIA behaved more indolent than LUAD. On the other hand, for down-trend patterns, natural killer mediated cytotoxicity and TGF-β signaling pathway were found to be decreased from normal lung tissue to AIS, suggesting an inhibited immune response against tumor. Chemokine signaling, IL2-STAT5 signaling, TNF-A signaling via NF-κB and leukocyte transendothelial migration were also found to be enriched in down-trend patterns, further suggesting an impaired immune function in tumor microenvironment. Interestingly, Wnt signaling pathway, which was often up-regulated in cancer, was found to be decreased from AIS to MIA and from MIA to LUAD. A deeper look into pattern 10 showed that this finding was contributed by WIF1 (Wnt inhibiting factor 1), which was an inhibitor of Wnt signaling pathway. Taken together, our results suggest that both an increased tumor intrinsic growth potential and an inhibited immune microenvironment contributed to the development and progression of lung adenocarcinoma.

### Decreased number of cytotoxic CD8+ T cells and increased number of Tregs in LUAD

Next, we used CIBERSORT, a deconvolution method to estimate the infiltration of immune cells in different pathological stages^34^. We found that the number of CD8+ T cells decreased as tumor stage increased (figure 3A), which was consistent with Dejima et al. on the immune evolution from neoplasia to invasive lung adenocarcionma^7^. Furthermore, we found that the number of Tregs increased as tumor stage increased, suggesting an enhanced immunosuppression as disease progressed (figure 3B), which was also validated by the Dejima dataset (figure 3C). Interestingly, we found an increased number of activated NK cells and a decreased number of resting NK cells as disease progressed, and validated this result using the Dejima dataset, suggesting NK cells might be important in the progression of lung adenocarcinoma (figure 3D and 3E). A more detailed staging of LUAD also showed similar results, with tumors at earlier stages having more CD8+ T cells and less Tregs, while tumors at later stages having less CD8+ T cells and more Tregs (supplementary figure 1). Taken together, our findings showed that a suppressed immune microenvironment plays an important role in the progression of lung adenocarcinoma.

**Figure 3.**
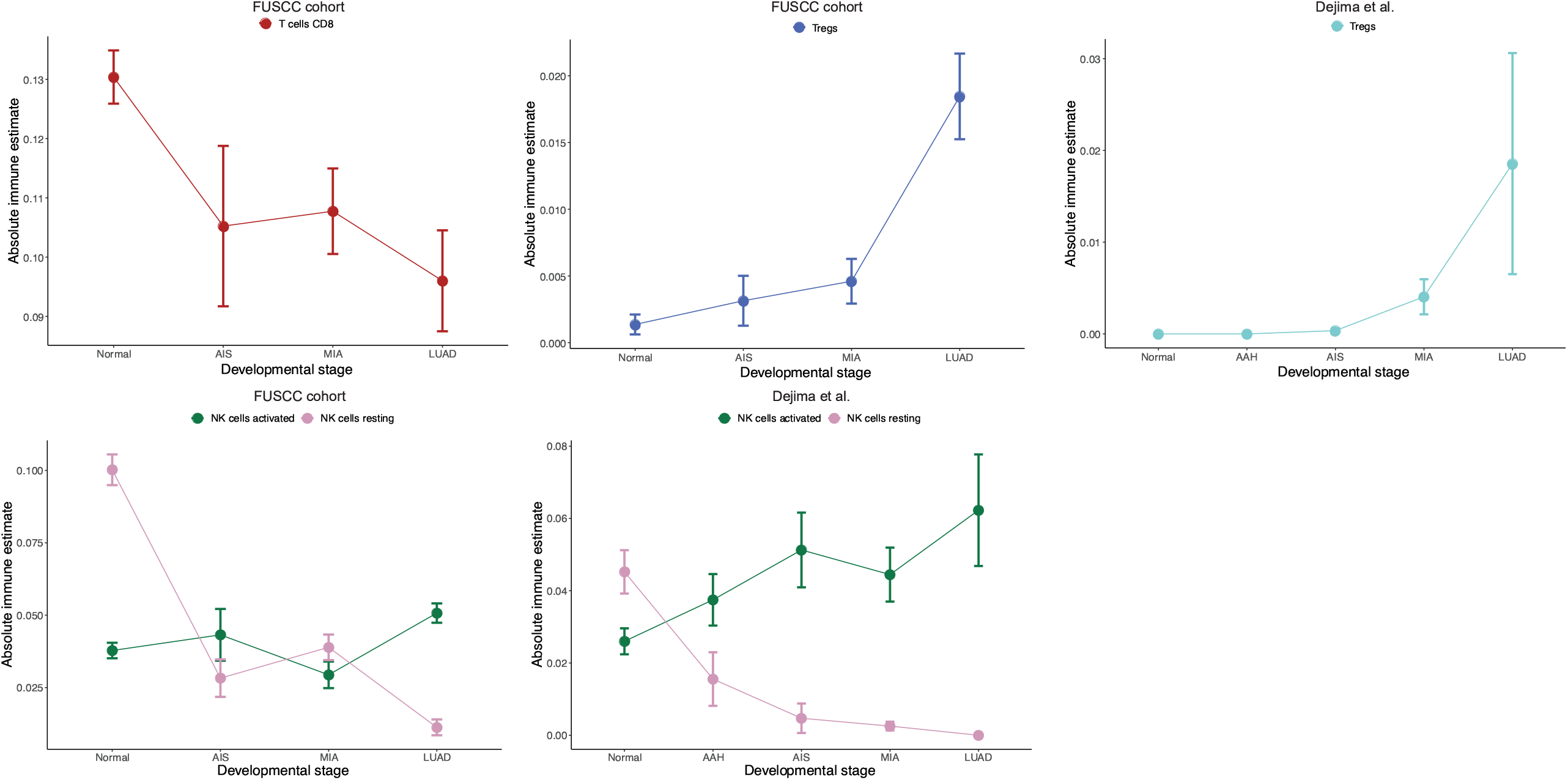
Immune cell infiltration prediction. A) Prediction of number of CD8+ T cells in the Fudan University Shanghai Cancer Center (FUSCC) cohort. B) Prediction of number of Tregs in the FUSCC cohort. C) Prediction of number of Tregs in Dejima’s cohort. D) Prediction of number of activated and resting natural killer (NK) cells in the FUSCC cohort. E) Prediction of number of activated and resting natural killer (NK) cells in Dejima’s cohort.

### Genomic alterations of tumors with different radiological manifestations

We next assessed the genomic alterations of tumors with different radiological manifestations. The landscape of somatic mutations for all patients included in this study was shown in Figure 4a. The most frequently mutated gene in this cohort was *EGFR* (50%), followed by *TP53* (22%), *RBM10* (8%), *ERBB2* (5%), *BRAF* (5%), *RB1* (5%), *KRAS* (4%) and *NF1* (3%), etc (figure 4A). A further comparison of mutation frequency in driver genes and tumor suppressor genes found that *BRAF* and *ERBB2* mutations were significantly enriched in pure GGOs and subsolid nodules (p = 0.046 and 0.016, respectively), while *ALK* fusions were detected only in solid nodules (3 out of 65, p = 0.045). There was no significant difference in total frequency of driver genes, however, a significant difference was observed in the frequency of tumor suppressor genes. The number of mutations in common tumor suppressor genes was 8 (11.6%) for the pure GGOs, 19 (30.2%) for subsolid nodules and sharply rose to 50 (76.9%) for solid nodules (p < 0.001, supplementary table 2). For individual tumor suppressor genes, frequency of *TP53* and *RB1* mutations was found to be significantly higher in solid nodules (p < 0.001 and p = 0.038, respectively, figure 4B, 4C and supplementary table 1). Tumor mutation burden was significantly higher in solid nodules compared to GGOs and subsolid nodules on CT scan (figure 4D).

**Figure 4.**
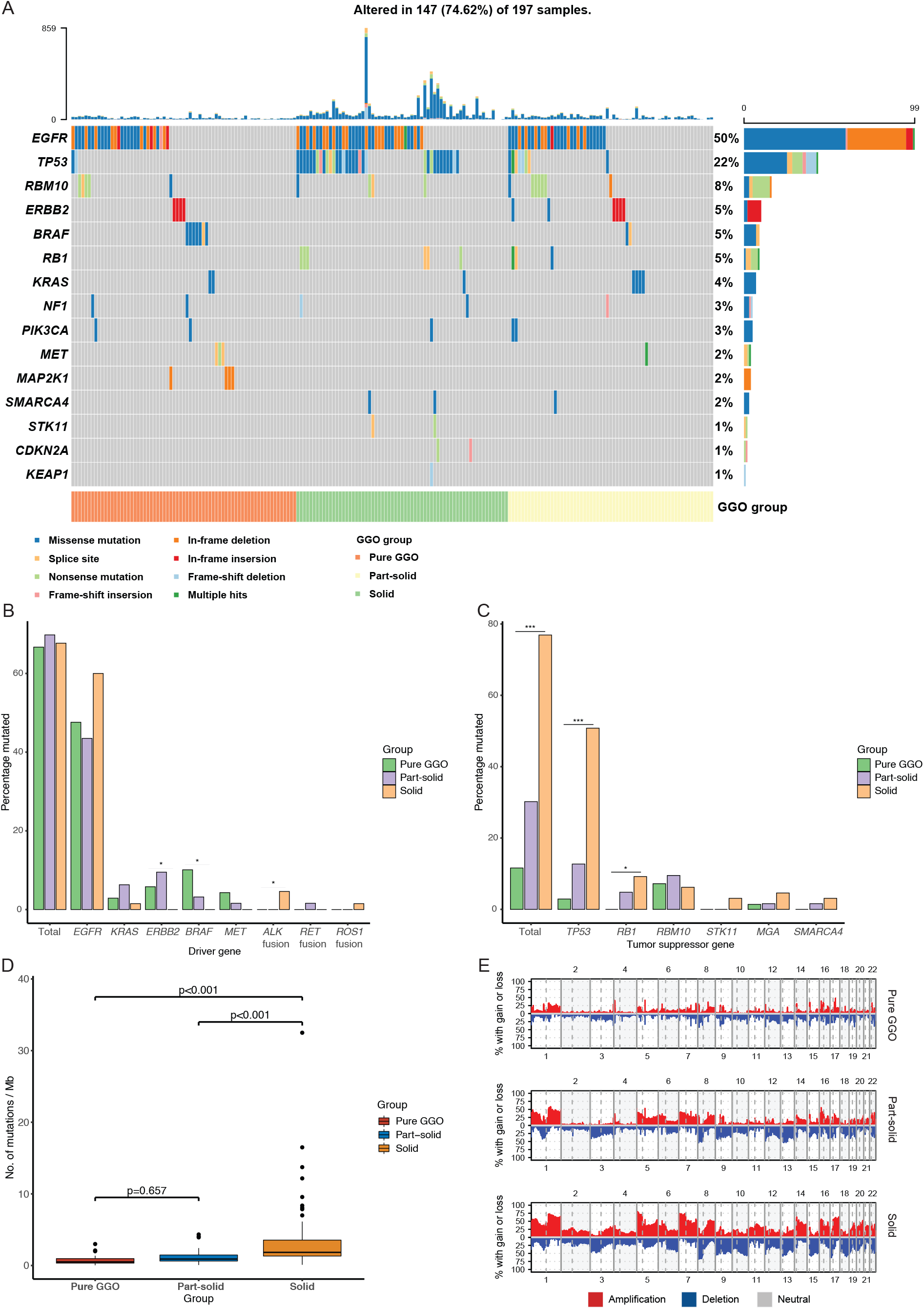
Comparison of genomic alterations for different radiological groups. A) Waterfall plot showing the landscape of genomic alterations in each radiological group. B) Comparison of mutation frequency in major driver genes among different radiological groups. C) Comparison of mutation frequency in tumor suppressor genes among different radiological groups. D) Comparison of tumor mutation burden among different radiological groups. E) Comparison of somatic copy number alterations among different radiological groups.

### Frequency of somatic copy number alterations were higher in solid nodules

To explore the somatic copy number alterations (SCNAs) of pure GGOs, subsolid and solid nodules, we identified SCNAs from raw sequencing reads of samples from the three groups. Notably, the SCNA frequency including amplification and deletion increased gradually from pure GGOs to solid nodules (figure 4E). We used GISTIC2.0 to identify significant arm-level SCNAs in samples of pure GGOs, subsolid and solid nodules. The union of significant arm-level event of the three groups was used for further analysis. As a result, the ratio of amplification and deletion in arm-level SCNAs also increased from pure GGOs to solid nodules. These results indicated that the frequency of somatic copy number alterations was positively associated with solid component in pulmonary nodules.

### Expression profile, differential expression analysis and gene set enrichment analysis (GSEA)

RNA-seq was performed to better understand the expression profiles of those radiologically different pulmonary nodules. Principle component analysis (PCA) showed that the transcriptomic profiles of the 3 groups were different (supplementary figure 2). Differential expression analysis demonstrated that 1359 genes were differentially expressed between the solid group and the GGO/subsolid group (|log2-fold change| >= 1, adjusted p value < 0.05, figure 5A and 4B). Of the 1359 differentially expressed genes, 870 were up-regulated and 489 were down-regulated (supplementary table 3). Furthermore, subgroup analyses showed that compared with the pure GGOs, subsolid nodules had 269 significantly up-regulated genes and 76 down-regulated genes, while compared with the subsolid group, the solid group had 668 significantly up-regulated genes and 320 down-regulated genes (supplementary figures 3-4 and supplementary table 4-5). There were 119 up-regulated genes and 20 down-regulated genes that were in common in the subgroup comparisons.

**Figure 5.**
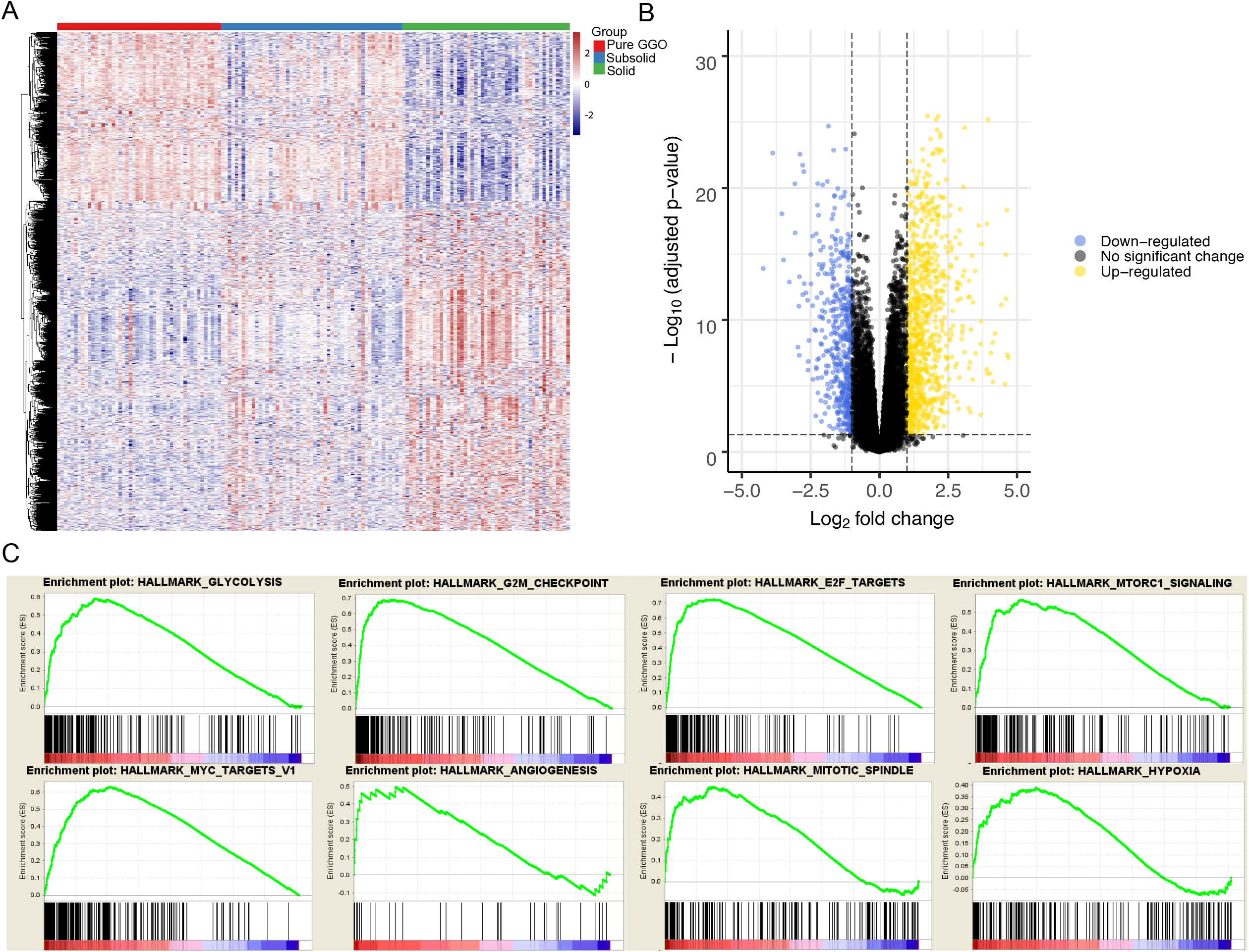
Differential expression analysis between the solid group and the pure GGO + subsolid group. A) Expression of differentially expressed genes between the solid group and the pure GGO + subsolid group. B) Volcano plot showing the number of significantly differentially expressed genes. C) Gene-set enrichment analysis (GSEA) showing the pathways that were enriched by differentially expressed genes.

To investigate the functional basis of the changes between GGOs/subsolid nodules and solid nodules, we performed gene set enrichment analysis (GSEA) on the RNA-seq data. Among all the predefined cancer hallmark pathways, genes associated with cellular proliferation and tumor growth were significantly enriched in the solid group, including glycolysis (NES=2.05, p < 0.001), G2M checkpoint (NES = 1.86, p = 0.006), E2F targets (NES = 1.84, p = 0.004), the MTORC1 signaling pathway (NES = 1.79, p = 0.004), MYC targets v1 (NES = 1.71, p = 0.012) and mitotic spindle (NES = 1.50, p = 0.048). Moreover, genes associated with angiogenesis (NES = 1.57, p = 0.041) and hypoxia (NES = 1.56, p = 0.031) were also found to be significantly enriched in the solid group (figure 5C). Taken together, these results suggest that compared with pure GGOs and subsolid nodules, solid nodules have a higher proliferation rate and tumor cells behave more malignantly.

### Tumor progressive index showed an imbalance between tumor intrinsic growth potential and immune response

Finally, we developed a tumor progressive index which quantitatively measures the level of imbalance between tumor intrinsic growth potential and immune response against tumors. Based on our previous findings, we used the genes in Pattern 1 and immune-related genes in Patterns 7 & 8 with mean TPM >= 1 across all samples (methods). 4 genes that were associated with apoptosis, tumor metastasis and progression (*BCL2L15, COMP, CST1* and *FAM83A*) reflected tumor intrinsic growth potential, while 8 genes (*ITLN2, MARCO, C8B, MASP1, CD36, TAL1, PPBP* and *CDH5*) that were associated with immune microenvironment reflected immune response against tumors. A negative tumor progressive index indicates that the immune system is competent enough to suppress the progression of tumors, while a positive tumor progressive index indicates that the immune system can no longer suppress the growth of tumor cells. In our study cohort, tumor progressive index was negative in normal tissues but positive in AIS and stages onwards, indicating immune escape already existed in AIS, the precursor stage of lung adenocarcinoma, and became more severe with the progression of disease (figure 6A). Same increasing trend was observed in another dataset which showed a significant increase of tumor progressive index from normal tissue to atypical adenomatous hyperplasia (AAH) then to LUAD (figure 6B)^35^. Interestingly, there was a negative tumor progressive index in the stage of AAH, where tumors could not overcome the immune system to metastasize further. Survival analyses showed that patients with a high tumor progressive index had poorer RFS and OS (figure 6C and 6D). Taken together, our results suggested that increased tumor intrinsic growth potential and impaired immune response against tumor work together to drive the progression of lung adenocarcinoma (figure 6E).

**Figure 6.**
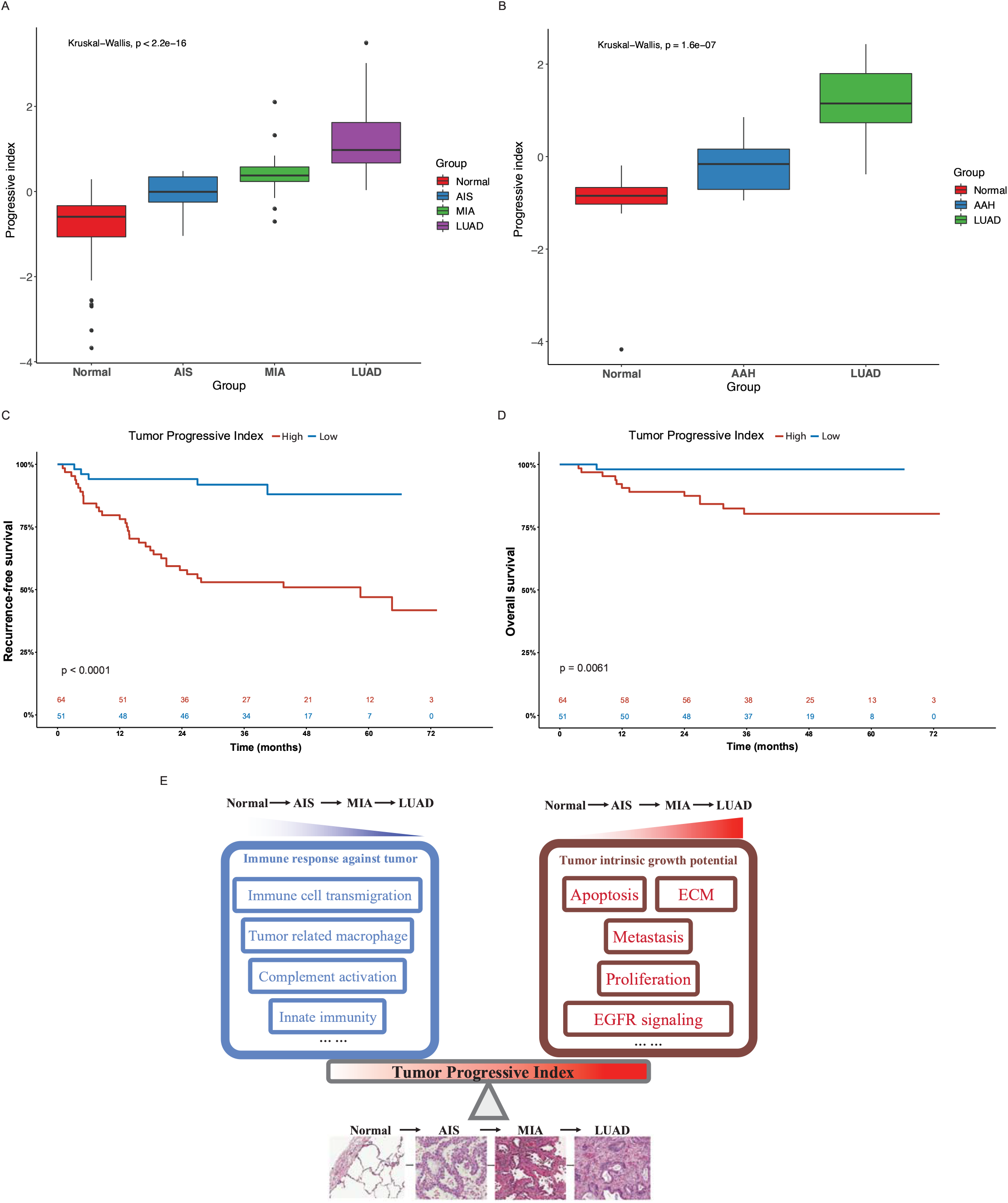
Comparison of tumor progressive index for different groups and the prognostic value of tumor progressive index. A) boxplot showing the index increased as tumor progressed in the FUSCC cohort. B) boxplot showing the index increased as tumor progressed in Dejima’s cohort. C) Recurrence-free survival for patients with solid vs subsolid and pure GGO/subsolid nodules. D) Overall survival for patients with solid vs subsolid and pure GGO/subsolid nodules.

## Discussions

In this study, we provided a comprehensive analysis integrating clinical, radiological, pathological, genomic and transcriptomic analysis of 197 pulmonary lesions with different radiological and pathological manifestations.

We first assessed the transcriptomic profiles of our cohort. Based on the differentially expressed genes between each two adjacent groups, 12 expression patterns were identified (figure 2). Genes associated with cell cycle and G2M checkpoint were found to be significantly up-regulated from MIA to LUAD, but not different from normal tissue to MIA. This indicates that pre-invasive stages of lung adenocarcinoma had more indolent behaviors, which might explain why AIS and MIA had a nearly 100% 5-year survival rate after surgical resection. Histologically, AIS is defined as a ≤3 cm adenocarcinoma lacking invasive patterns, while MIA is a ≤3 cm adenocarcinoma with invasive patterns of no more than 5 mm in size^33^. PI3K-AKT signaling pathway, a classical pathway that were up-regulated in various cancer types, were found to be increased from normal tissue to AIS^36^. Though AIS is the precursor of LUAD, this finding shows that cells in this stage already have an increased potential for growth. On the other hand, we found that epithelial-mesenchymal transition, a biological process known to increase metastatic potential of cancer cells^37^, were found to be significantly increased from AIS to MIA, consistent with histological differences between AIS and MIA. Though AIS and MIA both have a nearly perfect prognosis, our results suggest that MIA should be surgically intervened before it progresses to the next stage as invasive patterns already exist. To our surprise, several immune related pathways were found in down-trend expression patterns, indicating a decreased or impaired immune response as tumors progress. Consistent with this finding, using CIBERSORT to deconvolute our bulk-sequencing data, we found a decrease in the number of CD8+ cytotoxic T cells and an increase in the number of Treg cells, which was also validated by another dataset that compared immune cells among normal tissue, atypical adenomatous hyperplasia (AAH), AIS, MIA and LUAD^7^. Taken together, our results demonstrated that increased tumor intrinsic growth signals and decreased immune response orchestrate the evolution of lung adenocarcinoma.

We next demonstrated that there was an association between radiological and pathological presentations. Most nodules that were GGOs on CT scan were AIS or MIA, while most solid nodules were invasive lung adenocarcinoma (table 1). This might provide explanation of why pulmonary nodules manifesting as GGOs on CT scan usually have indolent clinical courses. This result was also consistent with previous studies and provided a link between histological and radiological manifestations^38,39^. For major driver mutations, we found that frequency of neither *EGFR* nor *KRAS* mutations was significantly different among the 3 groups (figure 4B and supplementary table 2). *BRAF* mutations were more enriched in the pure GGO group, while *ALK* fusion were only seen in the solid group. On the other hand, the frequency of mutations in tumor suppressor genes were significantly different among the 3 groups, with a sharp increase from 2 (2.9%) in the pure GGO group, to 8 (12.7%) in the subsolid group and 33 (50.8%) in the solid group (figure 4C and supplementary table 2), suggesting a pivotal role that tumor suppressor genes play in the progression of lung adenocarcinoma. For individual tumor suppressor genes, *TP53* and *RB1* were significantly different among the 3 groups (p < 0.001 and p = 0.038, respectively, fugure 3C and supplementary table 2). Interestingly, we incidentally found that there was a significant difference in *EGFR* mutation types among the 3 groups. *EGFR* exon 19 deletion was more enriched in the GGO group, while *EGFR* L858R was more often seen in the solid group (supplementary figure 5). Although in clinical settings, treatment for lung adenocarcinoma patients harboring *EGFR* L858R and *EGFR* exon 19 deletion is the same, our results indicates that those two major types of *EGFR* mutations might render tumors different malignant potentials.

Previous studies have discussed the genomic alterations in pulmonary nodules manifesting as GGO. In 2015, Kobayashi et al. evaluated *EGFR, KRAS, ALK* and *HER2* mutations in 104 pulmonary nodules manifesting as GGO. They found that *EGFR* was mutated in 64% of all the 104 samples, while *KRAS, ALK* and *HER2* were mutated in 4%, 3% and 4% of the samples, respectively^40^. In 2018, Lu et al. reported that *EGF*R was mutated in 75 out of 156 (48.1%) lung adenocarcinoma patients, which was similar to 50% in our cohort^41^. However, we did not observe a significant difference in *EGFR* mutation frequency among those 3 groups (figure 4B and supplementary table 2). In 2020, Li et al. performed whole-exome sequencing on 154 pulmonary subsolid nodules from 120 patients and found that *EGFR* were the most frequently mutated gene, followed by *RBM10, TP53, STK11* and *KRAS*. They also found that frequency of *EGFR, RBM10* and *TP53* was significantly different between pure GGO and subsolid nodules^42^. While not significantly different for *EGFR* and *RBM10* in our cohort, there was indeed a significant difference of the frequency of *TP53* mutations (figure 4C and supplementary table 2).

For somatic copy number alterations (SCNAs), we found that the frequency of SCNAs increased from pure GGO to subsolid and solid nodules. As the solid components increased, the genome became more unstable. Frequency of arm-level alterations also followed the same trend, where pure GGO had fewer significant arm-level events and solid nodules had more significant arm-level events. Genomic instability was widely reported to be associated with increased rate of tumor proliferation and progression among different cancer types^43–45^. Due to a higher level of genomic instability, solid nodules behave more malignant than GGO and subsolid nodules. Another unique advantage of our study is the application of RNA-seq in our cohort. By adding another aspect of analysis, we found that there were 1359 differentially expressed genes between the GGO/subsolid group and the solid group. GSEA analysis revealed that genes associated with cellular proliferation and tumor growth were significantly enriched in the solid group, including glycolysis, G2M checkpoint, E2F targets, the MTORC1 signaling pathway, MYC targets v1 and mitotic spindle, along with angiogenesis and hypoxia, which had a well-established role as a microenvironmental factor promoting tumor metastasis^46^.

Tumor intrinsic growth potential and immune microenvironment are both associated with tumor evolution and progression^47,48^. Based on our findings, a tumor progressive index was designed to quantitatively measure the level of imbalance between tumor intrinsic growth potential and immune microenvironment. Interestingly, AAH, a stage of pre-cancerous hyperplasia, had a negative tumor progressive index, indicating some key alteration would be needed to activate the malignant transformation of cells. AIS, MIA and LUAD all had a progressive index of more than 0, and the index increased as the tumor developed to the next stage. Furthermore, patients with a high tumor progressive index tended to have a poorer recurrence-free survival and overall survival. Taken together, tumor progressive index can be used as a predictor for disease progression and prognosis.

In summary, this study integrates the genomic alterations, transcriptomic profiles, and histological and radiological progression of lung adenocarcinoma, providing deeper understandings of the evolution of this disease.

## Data Availability

All data produced in the present study are available upon reasonable request to the authors.

## Acknowledgements

This work was supported by the National Natural Science Foundation of China (81930073), Shanghai Science and Technology Innovation Action Project (20JC1417200), Shanghai Municipal Science and Technology Major Project (2017SHZDZX01, VBH1323001/026), Shanghai Municipal Key Clinical Specialty Project (SHSLCZDZK02104) and Pilot Project of Fudan University (IDF159045).

## Authors’ contributions

Haiquan Chen, Leming Shi, Zhiwei Cao and Yue Zhao designed the study. Yue Zhao, Jun Shang and Jian Gao performed data analysis. Yue Zhao, Han Han and Zhendong Gao performed experiments. Qiang Zheng and Yuan Li reviewed pathological slides. Yueren Yan, Ting Ye, Fangqiu Fu, Chaoqiang Deng, Zelin Ma, Yang Zhang, Difan Zheng and Shanbo Zheng collected samples. Jun Shang and Leming Shi helped with quality control of sequencing data. Yue Zhao, Jun Shang and Jian Gao drafted the manuscript. All authors proofread the manuscript.

## Figure legends

Supplementary figure 1. Prediction of number of A) CD8+ T cells, B) Tregs and C) natural killer (NK) cells for different tumor stages.

Supplementary figure 2. Principle component analysis (PCA) of expressional profiles different pathological groups.

Supplementary figure 3. Differential analysis between solid and subsolid nodules. A) Expression of differentially expressed genes between solid and subsolid nodules. B) Volcano plot showing the number of significantly differentially expressed genes.

Supplementary figure 4. Differential analysis between GGOs and subsolid nodules. A) Expression of differentially expressed genes between solid and subsolid nodules. B) Volcano plot showing the number of significantly differentially expressed genes.

Supplementary figure 5. Comparison of *EGFR* mutation type for tumors with different radiological manifestations.

## Notes

### Competing Interest Statement

The authors have declared no competing interest.

### Author Declarations

This study was approved by the Committee for Ethical Review of Research (Fudan University Shanghai Cancer Center Institutional Review Board No. 090977-1).

