## Supplementary figures and images for "Up-regulated tumor intrinsic growth potential and decreased immune function orchestrate the evolution of lung adenocarcinoma"

### Supplementary figure 1

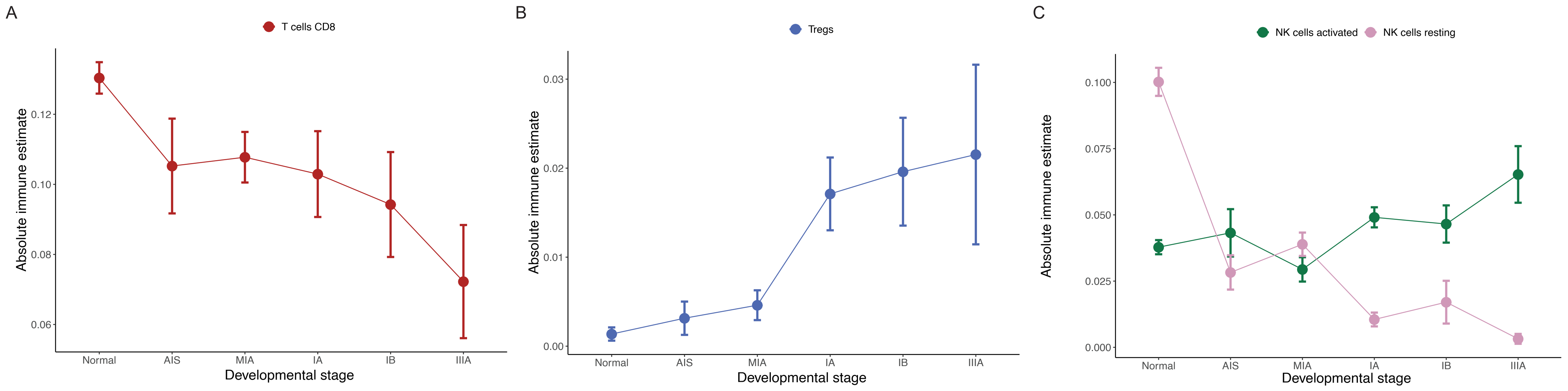

### Supplementary figure 2

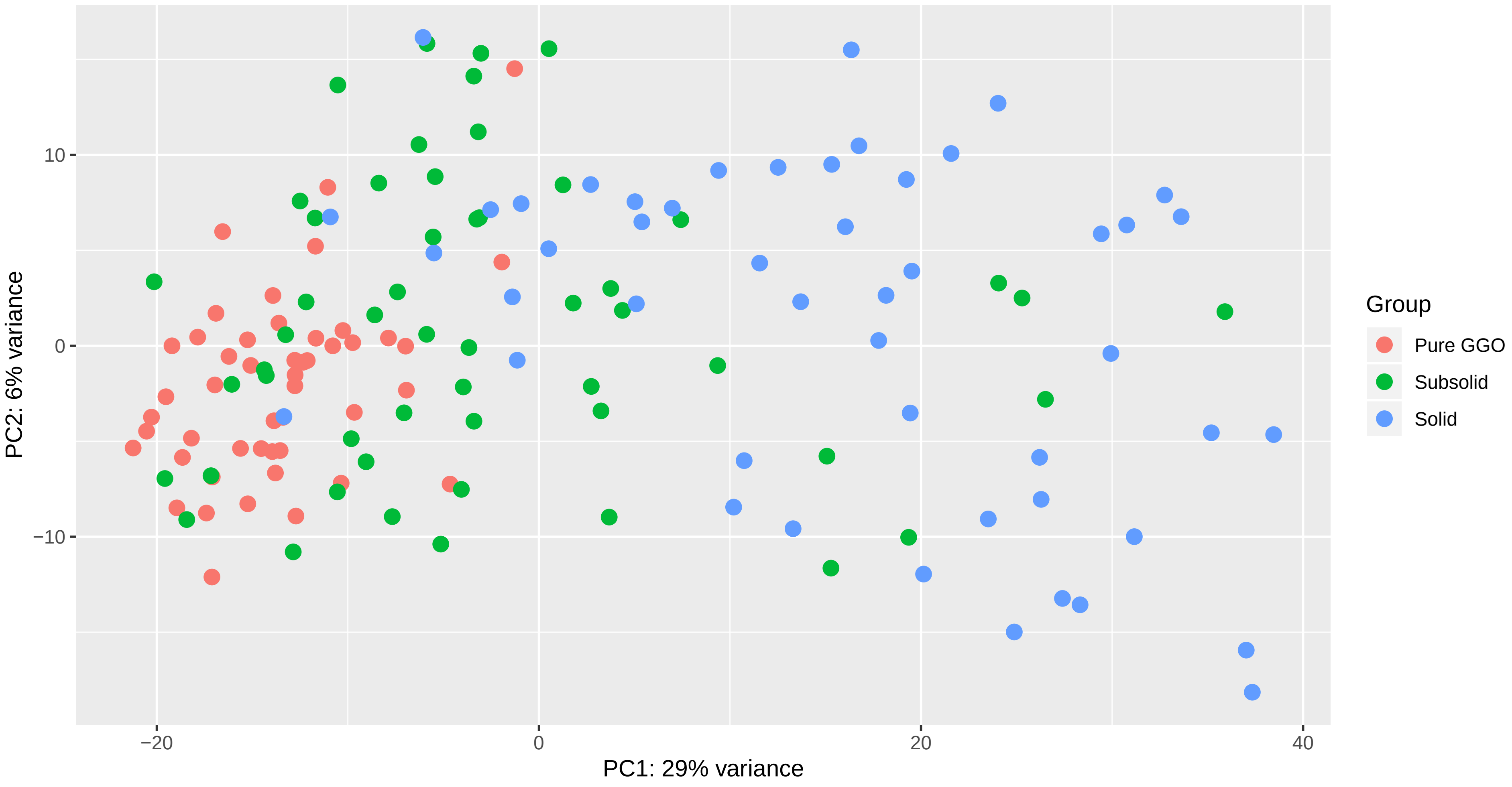

### Supplementary figure 3

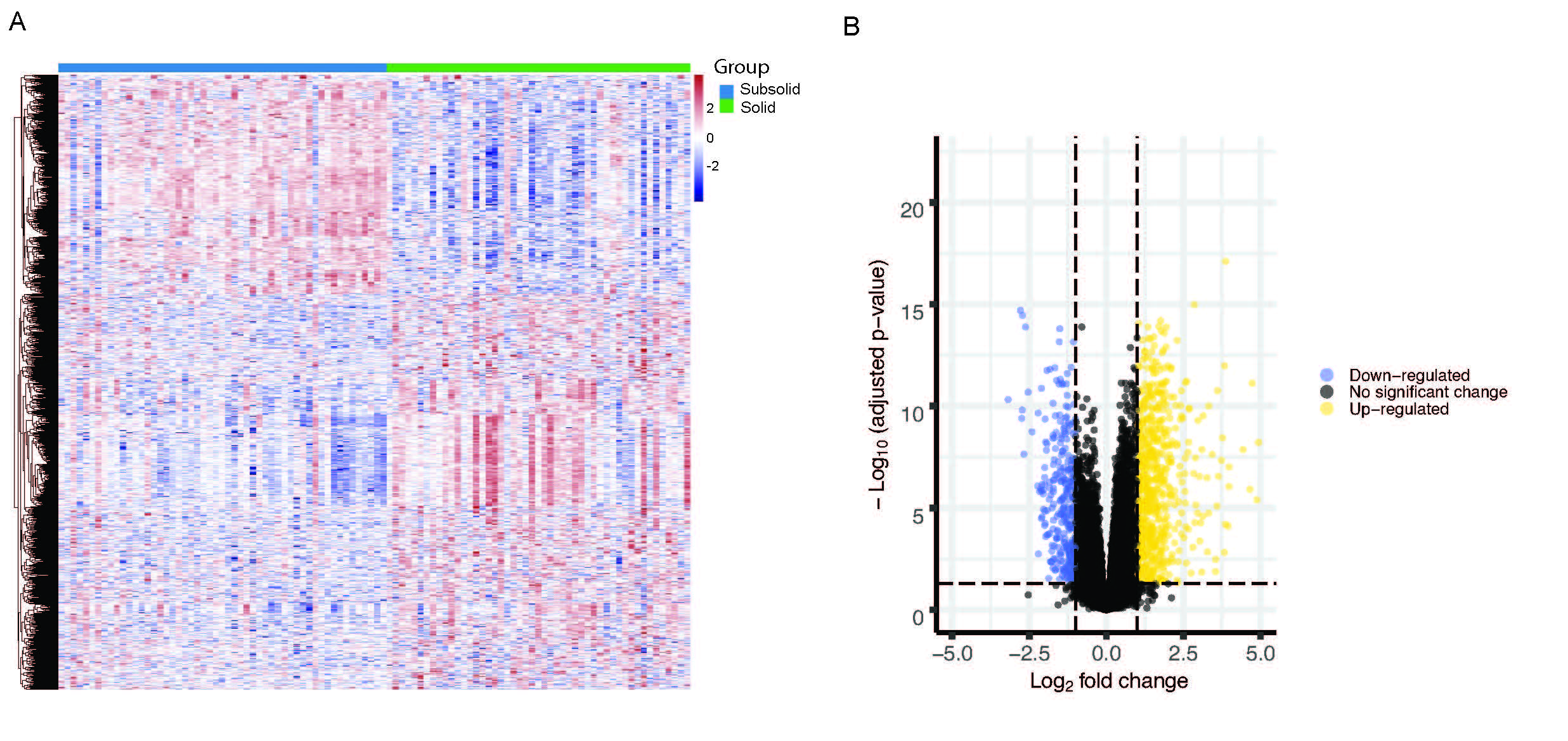

### Supplementary figure 4

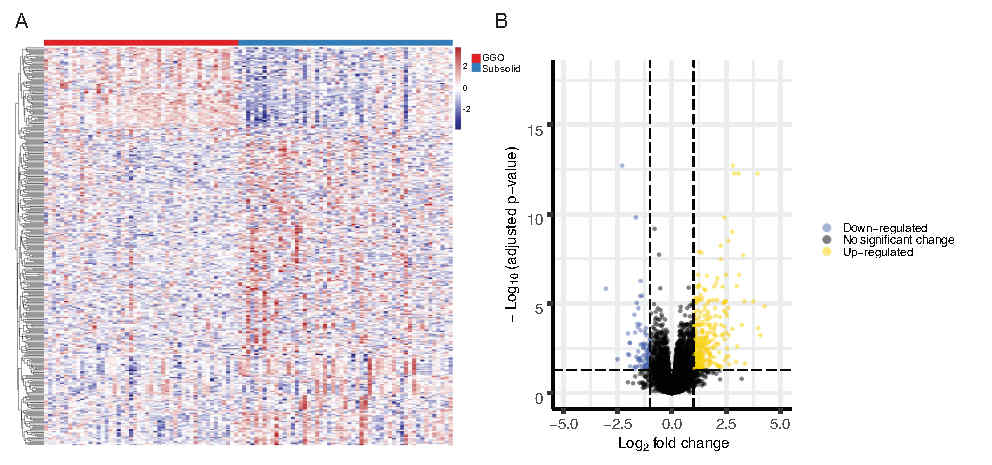
