## Supplementary figure 5 for "Up-regulated tumor intrinsic growth potential and decreased immune function orchestrate the evolution of lung adenocarcinoma"

| | Exon 19 Del | L858R | Exon 20 Ins | Others | P-value |
|---|---|---|---|---|---|
| Pure GGO | 17 (68.0%) | 1 (2.3%) | 0 (0.0%) | 4 (30.8%) | < 0.001 |
| Subsolid | 8 (32.0%) | 18 (41.9%) | 2 (66.7%) | 5 (38.5%) | |
| Solid | 0 (0.0%) | 24 (55.8%) | 1 (33.3%) | 4 (30.8%) | |
