## Supplementary table 2 for "Up-regulated tumor intrinsic growth potential and decreased immune function orchestrate the evolution of lung adenocarcinoma"

|  | Pure GGO  (n = 69) | Part-solid  (n = 63) | Solid  (n = 65) | P-value |
| --- | --- | --- | --- | --- |
| Driver genes | 46 (66.7%) | 44 (69.8%) | 44 (67.7%) | 0.989 |
| *EGFR* | 30 (47.6) | 30 (43.5) | 39 (60.0) | 0.141 |
| *KRAS* | 2 (2.9%) | 4 (6.3%) | 1 (1.5%) | 0.318 |
| *ERBB2* | 4 (5.8) | 6 (9.5) | 0 (0.0) | 0.046 |
| *BRAF* | 7 (10.1) | 2 (3.2) | 0 (0.0) | 0.016 |
| *MET* | 3 (4.3) | 1 (1.6) | 0 (0.0) | 0.195 |
| *ALK* fusion | 0 (0.0) | 0 (0.0) | 3 (4.6) | 0.045 |
| *RET* fusion | 0 (0.0) | 1 (1.6) | 0 (0.0) | 0.343 |
| *ROS1* fusion | 0 (0.0) | 0 (0.0) | 1 (1.5) | 0.360 |
| Tumor suppressor genes | 8 (11.6) | 19 (30.2) | 50 (76.9) | < 0.001 |
| *TP53* | 2 (2.9) | 8 (12.7) | 33 (50.8) | < 0.001 |
| *RB1* | 0 (0.0) | 3 (4.8) | 6 (9.2) | 0.038 |
| *RBM10* | 5 (7.2) | 6 (9.5) | 4 (6.2) | 0.765 |
| *STK11* | 0 (0.0) | 0 (0.0) | 2 (3.1) | 0.129 |
| *MGA* | 1 (1.4) | 1 (1.6) | 3 (4.6) | 0.429 |
| *SMARCA4* | 0 (0.0) | 1 (1.6) | 2 (3.1) | 0.347 |

Supplementary table 1. Mutational status of major driver and tumor suppressor genes in the study cohort (n = 197).
